# Patient and physician shared decision making behaviors in oncology: Evidence on adequate measurement properties of the iSHARE questionnaires

**DOI:** 10.1101/2021.02.12.21251610

**Authors:** Hanna Bomhof-Roordink, Anne M. Stiggelbout, Fania R. Gärtner, Johanneke E.A. Portielje, Cor D. de Kroon, Koen C.M.J. Peeters, Karen J. Neelis, Jan Willem T. Dekker, Trudy van der Weijden, Arwen H. Pieterse, for the iSHARE study group

## Abstract

**Objectives:** We have developed two questionnaires to assess the shared decision making (SDM) process in oncology; the iSHAREpatient and iSHAREphysician. In this study, we aimed to determine: scores, construct validity, test-retest agreement (iSHAREpatient), and inter-rater (iSHAREpatient-iSHAREphysician) agreement.

**Methods:** Physicians from seven Dutch hospitals recruited cancer patients, and completed the iSHAREphysician and SDM-Questionnaire–physician version. Their patients completed the: iSHAREpatient, 9-item SDM-Questionnaire, Decisional Conflict Scale, Combined Outcome Measure for Risk communication And treatment Decision making Effectiveness, and Perceived Efficacy in Patient-Physician Interactions. We formulated, respectively, one (iSHAREphysician) and 10 (iSHAREpatient) a priori hypotheses regarding correlations between the iSHARE questionnaires and questionnaires assessing related constructs. To assess test-retest agreement patients completed the iSHAREpatient again 1-2 weeks later.

**Results:** In total, 151 treatment decision making processes with unique patients were rated. Dimension and total iSHARE scores were high both in patients and physicians. The hypothesis on the iSHAREphysician and 9/10 hypotheses on the iSHAREpatient were confirmed. Test-retest and inter-rater agreement were >.60 for most items.

**Conclusions:** The iSHARE questionnaires show high scores, have good construct validity, substantial test-retest agreement, and moderate inter-rater agreement.

**Practice implications:** Results from the iSHARE questionnaires can inform both physician- and patient-directed efforts to improve SDM in clinical practice.

## 1. Introduction

*Those who have not experienced the intricacies of clinical practice demand measures that are easy, precise, and complete—as if a sack of potatoes was being weighed. True, some elements in the quality of care are easy to define and measure, but there are also profundities that still elude us. We must not allow anyone to belittle or ignore them; they are the secret and glory of our art*.

*Avedis Donabedian*[1]

Measurement of shared decision making (SDM) remains a challenge [2–4]. The SDM process in which patients, loved ones and healthcare professionals together arrive at treatment decisions incorporating patients’ values and preferences is not easy to capture in a measurement instrument. SDM happens both during and outside consultations [5], involves both observable (e.g., information-giving) and covert (e.g., thinking about the options) behaviors, and includes behaviors of both patients and healthcare professionals [6, 7]. Current SDM measurement instruments do not cover all of these aspects, and they substantially differ in which SDM elements are assessed [8, 9]. Many often-used measurement instruments assess only healthcare professional behavior (e.g., OPTION [10], CollaboRATE [11]) or do not assess patient behavior independently of physician behavior (e.g., 9-item SDM-Questionnaire (SDM-Q-9) [12], SDM-Questionnaire – physician version (SDM-Q-Doc) [13]), impeding the assessment of patients’ role.

We developed the iSHARE questionnaires to assess SDM in oncology, from both a patient (iSHAREpatient) and physician (iSHAREphysician) viewpoint [14]. We chose the oncology setting since cancer patients often face preference-sensitive decisions [15, 16]. The SDM construct was informed by an SDM model in oncology based on stakeholders’ views, and by a review of SDM models across healthcare settings published until June 2016. The iSHARE questionnaires include both patient and physician behaviors. Cancer patients and physicians were extensively involved during the development process, in line with quality criteria for the development of health-related measurement instruments [17].

The present study aimed to a) describe scores obtained by the iSHARE questionnaires in an oncology setting, and determine b) construct validity of the iSHARE questionnaires, c) test-retest agreement of the iSHAREpatient, and d) agreement between scores by the iSHAREpatient and iSHAREphysician.

## 2. Methods

### 2.1 Study design

In this multicenter study, we asked physicians from seven Dutch hospitals to complete a questionnaire after each consultation with an unique eligible patient, between June 2018 and December 2019. Participating patients were asked to complete a questionnaire after the consultation, and again after an aimed period of 1-2 weeks. The Medical Ethical Committee of the Leiden University Medical Center (LUMC) approved the study (NL50551.058.14, P14.207), which was conducted according to the Dutch Medical Research Involving Human Subjects Act.

### 2.2 Participant recruitment

We approached physicians treating cancer patients for participation, and asked consenting physicians to recruit consecutive unique eligible patients. Patients were eligible for the study if they had been diagnosed with cancer, were ≥18 years old, able to speak and write Dutch, had a consultation in which a decision to start, stop, change or forgo treatment with curative or palliative intent was discussed, and a life expectancy of over three months. We aimed to assess the measurement properties of the iSHARE questionnaires in a sample representing the heterogeneity of cancer treatment decisions, and therefore asked physicians from a range of cancer specialties to approach patients.

The physicians provided patients with an information letter, an informed consent form, and a post-consultation questionnaire, and asked them if they agreed to being called by the researchers. If so, we contacted them to ask if they had questions and if they were willing to participate. Consenting patients sent us their signed informed consent form and the completed questionnaire. We used the physician’s questionnaire if the patient had provided informed consent.

### 2.3 Data collection

Physicians reported their birth year, gender, year of start of specialization, working place, and specialty. They completed the iSHAREphysician [14] and the SDM-Q-Doc [13] post-consultation on paper or online. They also reported the patient’s primary tumor type and curative/palliative intent of the treatment discussed. Patients completed the: iSHAREpatient [14], SDM-Q-9 [12], Decisional Conflict Scale (DCS) [18], Combined Outcome Measure for Risk communication And treatment Decision making Effectiveness (COMRADE) [19], five-item Perceived Efficacy in Patient-Physician Interactions (PEPPI-5) [20], and birth date, gender, education, month and year of most recent cancer diagnosis, and number of consultations they had in mind while completing the questionnaire, also on paper or online. We sent consenting patients the iSHAREpatient again on paper or online, whichever they preferred, within a few days after we had received the initial questionnaire. We entered the data from the paper questionnaires in the online database questionnaire system Qualtrics.

### 2.4 iSHAREpatient and iSHAREphysician

The iSHAREpatient (Box 1) and iSHAREphysician (Box 2) each have the same, but mirrored 15 items [14], with a six-point unbalanced scale, ranging from ‘not at all’ (0) to ‘completely’ (5) [21]. They encompass the same SDM construct, consisting of six dimensions. The items relate to these six dimensions, which we do not assume to be necessarily correlated [2, 22, 23], leading us to adopt a formative measurement model (i.e., the items *form* the construct) [14]. The dimensions aim to assess the complete SDM process both during and outside consultations, and include both patient and physician behaviors. Depending on whether a decision has already been made or not, either item 15 or item 16 is relevant [14]. If a patient or physician had indicated that a decision had been made, or if the response to that item was missing, we report the score on item 15; otherwise, we report the score on item 16. The score on dimension six therefore consists of either the score on item 15 or 16.

We calculated dimension scores (range, 0-5) and a total score (the sum of the dimension scores; range, 0-30) for both iSHARE questionnaires. We applied a linear transformation to obtain a 0-100 total score ((score/30)*100). Higher dimension and total scores indicate higher levels of SDM. We only report dimension and total scores if all the respective items had been completed; the formative nature of the construct makes imputation of missing values inappropriate.

### 2.5 Construct validity of the iSHAREpatient and iSHAREphysician

We determined construct validity by testing hypotheses about correlations between the iSHARE questionnaires and questionnaires measuring related constructs. We formulated a priori hypotheses on total score level for both iSHARE questionnaires, and on dimension and item level for the iSHAREpatient (Table 5). We further expected the three iSHAREpatient items on patient-initiated behavior (items 7, 13 and 14) each to correlate with the PEPPI-5. Based on the respective content of the scales and items we expected a correlation of >.30 or <-.30 for each hypothesis.

**Table 5.** Correlations between the iSHARE and other questionnaires

| iSHARE questionnaire | Comparison scale - subscale | N | Spearman Rho* |
| --- | --- | --- | --- |
| iSHAREphysician | SDM-Q-Doc | 155 | .84~ |
| iSHAREpatient | SDM-Q-9 | 144 | .77~ |
|  | COMRADE – Satisfaction with communication | 125 | .68~ |
| iSHAREpatient dimension (item) |  |  |  |
| 2. Medical information (1-7) | DCS – Feeling uninformed | 146 | -.44~ |
| 2. Medical information (7)<sup>a</sup> | PEPPI-5 | 154 | .31~ |
| 3. Preferences (10,13) | DCS – Feeling unclear about values | 151 | -.43~ |
| 3. Preferences (13)<sup>a</sup> | PEPPI-5 | 155 | .40~ |
| 4. Deliberation (14)<sup>a</sup> | PEPPI-5 | 154 | .27 |
| 6. Decision (15) | DCS – Ineffective decision making | 138^ | -.46~ |
| 6. Decision (16) | DCS – Feeling unsupported | 15^ | -.66~ |

*Note.* Hypotheses were formulated a priori on total score level for both iSHARE questionnaires, and on dimension and item level for the iSHAREpatient. The expected correlation was >.30 for the SDM-Q-9, SDM-Q-Doc, COMRADE and PEPPI-5 and <-.30 for the DCS. No a priori hypotheses were formulated for the iSHAREphysician dimensions, and neither for three iSHAREpatient dimensions (i.e., 1. Choice Awareness, 4. Deliberation and 5. Time for Deliberation).
<sup>^</sup>We report item 15 if a patient had reported that a decision had been made or if the response to that item was missing; we report item 16 if a patient had reported that no decision had been made.
\* $p < .01$
<sup>a</sup> Items measuring patient behavior
~Hypothesis was confirmed
COMRADE = Combined Outcome Measure for Risk communication And treatment Decision making Effectiveness; DCS = Decisional Conflict Scale; IQR = interquartile range; PEPPI-5 = five-item Perceived Efficacy in Patient-Physician Interactions; SDM-Q-9 = 9-item SDM-Questionnaire; SDM-Q-Doc = SDM-Questionnaire – physician version

#### 2.5.1 SDM-Q-9 and SDM-Q-Doc

The SDM-Q-9 [12] and SDM-Q-Doc [13] assess SDM from respectively patient and physician perspective. They each include nine items that are scored on a six-point scale from ‘completely disagree’ (0) to ‘completely agree’ (5). The raw score ranges from 0-45 and is multiplied by 20/9, resulting in a score from 0-100. Higher scores indicate higher levels of SDM [12, 13]. Both questionnaires have been validated in the oncology setting [24–26], and have been translated and validated in Dutch [27]. Cronbach’s α’s were .90 (SDM-Q-9) and .85 (SDM-Q-Doc).

#### 2.5.2 COMRADE

The COMRADE aims to measure effectiveness of risk communication and treatment decision making in consultations, and consists of two subscales: satisfaction with communication (10 items) and confidence in decision (10 items). The response scale ranges from ‘strongly disagree’ (1) to ‘strongly agree’ (5) [19]. We calculated subscale scores based on the original factor analysis that was provided by the developer. Both subscale scores range from 0-100, with higher scores indicating more satisfaction or confidence, respectively. The COMRADE has been translated in Dutch [28]. Cronbach’s α’s were .91 (satisfaction with communication) and .90 (confidence in decision).

#### 2.5.3 DCS

The DCS is a 16-item questionnaire assessing the level of decisional conflict; the five-point scale items range from ‘strongly agree’ (0) to ‘strongly disagree’ (4) [18]. The scale consists of five subscales: feeling uncertain (3 items), feeling uninformed (3 items), feeling unclear about values (3 items), feeling unsupported (3 items), and ineffective decision making (4 items) [29]. To calculate the subscale scores, item scores are summed, divided by the number of items in the subscales and multiplied by 25, with scores ranging from 0 to 100. The total score ranges from 0-64, is multiplied by 25/16, resulting in a standardized score from 0 to 100. Higher scores indicate higher decisional conflict. The DCS has been translated and validated in Dutch, in an oncology setting [30]. Cronbach’s α’s were .69 (feeling uncertain), .73 (feeling uninformed), .58 (feeling unclear about values), .32 (feeling unsupported) and .82 (ineffective decision making).

#### 2.5.4 PEPPI-5

The PEPPI-5 aims to measure patients’ perceived self-efficacy in obtaining medical information and attention to their medical concerns from physicians. The response scale ranges from ‘not at all confident’ (1) to ‘very confident’ (5) and the total score ranges from 5 to 25, with higher scores representing higher perceived self-efficacy in patient-physician interactions [20]. The PEPPI-5 has been translated and validated in Dutch, in patients with osteoarthritis [31]. Cronbach’s α was .91.

### 2.6 Test-retest agreement of the iSHAREpatient

We assessed test-retest agreement of the iSHAREpatient, that is, the extent to which item scores for patients with a stable perception of the SDM process were the same for repeated measurements over time [32]. The COnsensus-based Standards for the selection of health Measurement Instruments (COSMIN) study design checklist [33] requires participants to be stable during the chosen interval, and the interval to be long enough to avoid them recalling their scores at first administration; we expected a time window of 1-2 weeks to be appropriate between test and retest. We excluded patients who answered affirmatively to one or both of the following questions at retest: ‘Please think back to the time you filled in the questionnaire for the first time. Do you have different thoughts regarding the decision making process now, compared to the thoughts you had back then? ’ and ‘Have you had another conversation with the physician in the meantime? ’.

We did not consider it feasible to assess test-retest agreement for the iSHAREphysician. We did not expect physicians to be able to recall the treatment decision making process for a particular patient well enough over a period of 1-2 weeks to complete the iSHAREphysician again for that patient.

### 2.7 Inter-rater agreement between the iSHAREpatient and iSHAREphysician

In accordance with the COSMIN [33] study design checklist we determined agreement (not correlation) between the scores on the iSHAREpatient and iSHAREphysician.

### 2.8 Statistical analyses

#### 2.8.1 Selection and missing values

We excluded patient questionnaires if they had been completed >30 days post-consultation and physician questionnaires if they had been completed >7 days post-consultation (Figure 1). We assumed that a longer period would be detrimental to participants’ recollection of the decision making process.

**Figure 1.**
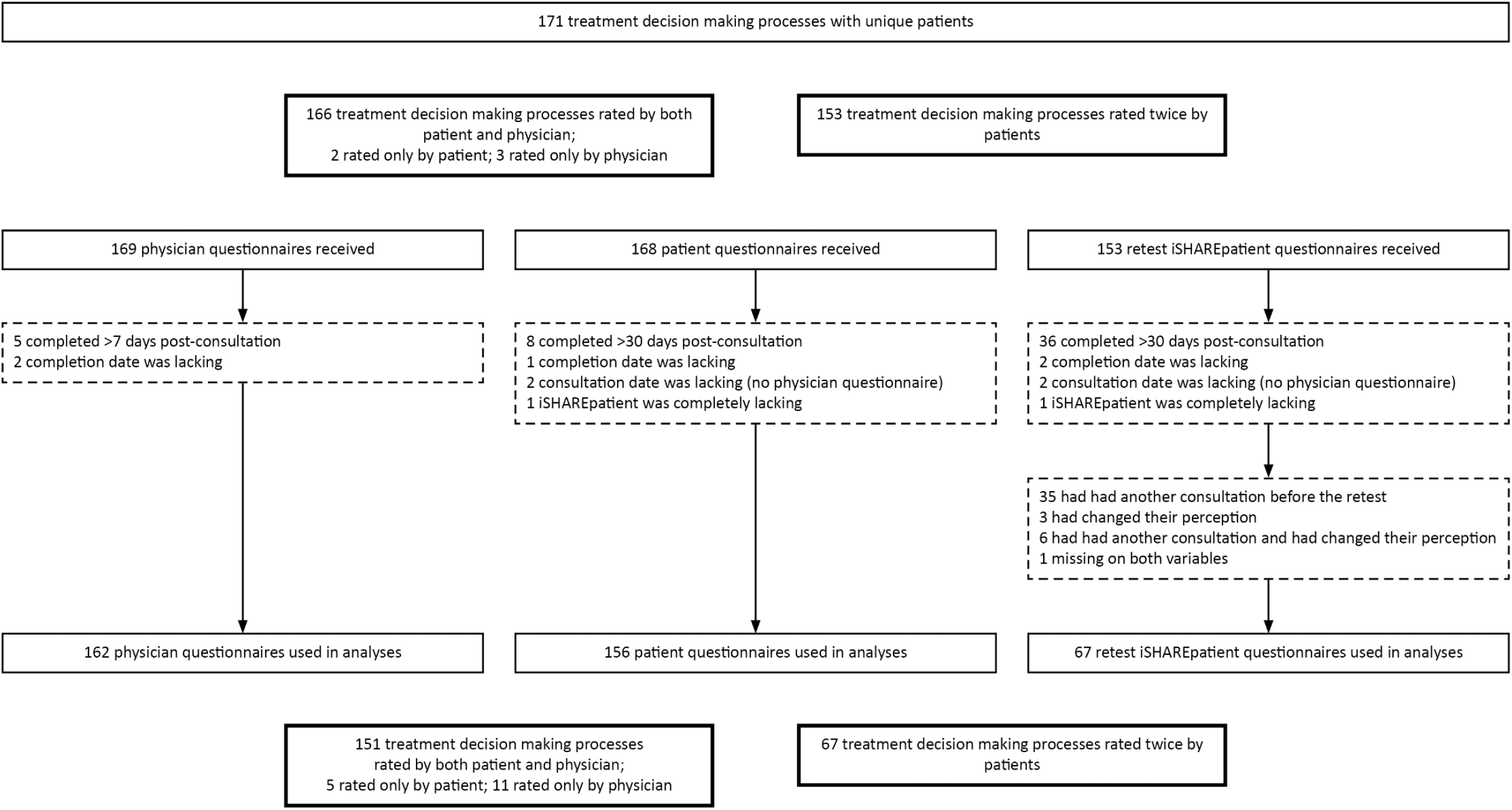
Flow diagram of participants

We handled missing values according to authors’ recommendations, if provided in the original or Dutch validation paper (see section 2.5) [12, 13, 31]. For the other questionnaires and the iSHARE questionnaires (see section 2.4), we only report scores when all respective items had been completed. We report sample sizes per analysis, since these may differ due to missing values.

#### 2.8.2 Analyses

Descriptive statistics were used to report scores on all questionnaires. Hypotheses were tested by calculating Spearman correlation coefficients between the scores on the iSHARE questionnaires and the respective comparison questionnaires, as the data were non-normally distributed on all scales. We determined test-retest agreement and inter-rater agreement by calculating agreement and the corresponding 95% confidence intervals (CIs) [34, 35]. Due to the non-normally distributed data it was not possible to calculate weighted kappa’s. For test-retest agreement we defined agreement as the same item score obtained both at test and retest: (X_00_+X_11_+X_22_+X_33_+X_44_+X_55_)/(X_01_+X_02_+X_03_+X_04_+X_05_+X_10_+X_12_+…+X_54_), where e.g., X_33_ means that for both test and retest the item score was 3. For inter-rater agreement, we allowed the item scores to differ one point, since we considered it acceptable if scores from the respective viewpoints somewhat differed. To illustrate, a score of 5 on an iSHAREpatient item and a score of 4 on the same iSHAREphysician item (i.e., X_54_), was considered as agreement. Consequently, agreement was defined as:

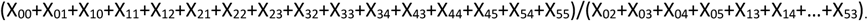

The corresponding CIs were calculated as follows:

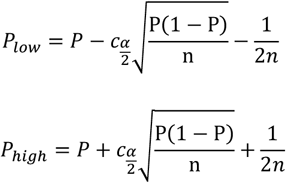

When the lower limit was ≤.3 or the upper limit ≥.7, the Fleiss correction was applied. These limits were calculated as follows [34]:

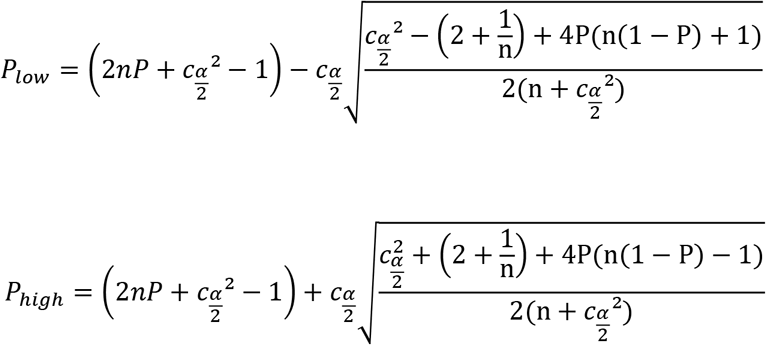

where *P* is the proportion agreement, *n* the sample size and 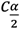 the percentile cut-off for the standard normal distribution (i.e., 1.96 for the 95% CI). Confidence intervals for agreement were calculated in Excel version 2010. We used SPSS version 25 to perform all other analyses. A *p*-value <.05 was considered statistically significant.

## 3. Results

### 3.1 Participants

In total, 156 patients and 51 physicians participated in the study (Table 1). Fifty-seven eligible patients who had been approached for participation by their treating physician and took the study information home, did not provide consent. We do not know how many eligible patients have been approached and declined immediately. In total, 151 treatment decision making processes were rated by both patients and physicians, with a range of one to seven per physician. Five decision processes were only rated by patients and eleven only by physicians (Figure 1). Patients completed the initial questionnaire 6.0±6.0 (range, 0-29) days post-consultation and physicians 0.2±0.8 (range, 0-7) days post-consultation. Eighty-five patients thought about more than one consultation while completing the questionnaire.

**Table 1.** Patient (n=156) and physician (n=51, who rated 162 treatment decision making processes) socio-demographic, and disease- or work-related characteristics

| | N* | Percentage or mean ± SD |
| --- | --- | --- |
| **Patients** |  |  |
| Sex, female | 67 | 43% |
| Age, years | 156 | 67.5 ± 12.5 |
| Education level | 153 |  |
| *Low* | 46 | 30% |
| *Intermediate* | 43 | 28% |
| *High* | 64 | 42% |
| Primary tumor type | 156 |  |
| *Gastro-intestinal* | 42 | 27% |
| *Urological* | 36 | 23% |
| *Breast* | 22 | 14% |
| *Lung* | 17 | 11% |
| *Hematological* | 13 | 8% |
| *Gynecological* | 10 | 6% |
| *Other* | 16 | 11% |
| Treatment intent | 154 |  |
| *Curative* | 90 | 58% |
| *Palliative* | 59 | 38% |
| *Other* | 5 | 3% |
| Months since most recent cancer diagnosis | 143 |  |
| *0-3* | 66 | 46% |
| *4-12* | 34 | 24% |
| *>12* | 43 | 30% |
| **Physicians** |  |  |
| Sex, female | 24 | 47% |
| Age, years | 51 | 44.4 ± 9.6 |
| Years since start specialist training | 51 | 15.8 ± 8.4 |
| Hospital | 52 |  |
| *Academic (n=2)* | 33 | 65% |
| *Non-academic (n=5)* | 18 | 35% |
| *Table 1 continued* |  |  |
| Specialty | 51 |  |
| *Radiotherapy* | 17 | 33% |
| *Medical Oncology* | 11 | 22% |
| *Urology* | 6 | 12% |
| *Surgery* | 4 | 8% |
| *Gynecology* | 3 | 6% |
| *Pulmonology* | 4 | 8% |
| *Other* | 6 | 12% |

\*Numbers do not always add up to the total sample size, due to missing values.
SD=Standard deviation

### 3.2 Responses on the iSHAREpatient and iSHAREphysician

Both the iSHAREpatient and iSHAREphysician showed few missing values (Table 2). The iSHAREpatient and iSHAREphysician dimension scores showed a distribution skewed toward higher scores (Figure 2). Median total scores (interquartile range (IQR)) were 95.0 (77.1-99.5) (iSHAREpatient) and 75.0 (61.1-90.7) (iSHAREphysician) (Table 3). In total, 35 (23%) patients and for 15 (10%) treatment decision making processes physicians gave the highest possible total score (100).

**Table 2.** Median, interquartile range, range, and agreement on item level for the iSHAREpatient (n=156 treatment decision making processes) and iSHAREphysician (n=162 treatment decision making processes)

|  | iSHAREpatient |  | iSHAREphysician |  | Agreement~ |  |
| --- | --- | --- | --- | --- | --- | --- |
|  | N | Median (IQR), range | N | Median (IQR), range | N | P (95%CI) |
| *Item score (0-5)* |  |  |  |  |  |  |
| 1. Physician explained advantages of treatment options | 155 | 5.0 (4.0-5.0), 0-5 | 162 | 4.0 (4.0-5.0), 0-5 | 150 | .79 (.71* - .84*) |
| 2. Physician explained disadvantages of treatment options | 156 | 5.0 (3.0-5.0), 0-5 | 162 | 4.0 (4.0-5.0), 2-5 | 151 | .72 (.64 - .78*) |
| 3. Physician explained (dis)advantages equally well | 154 | 5.0 (3.0-5.0), 0-5 | 161 | 4.0 (3.0-5.0), 0-5 | 148 | .68 (.60 - .75*) |
| 4. Physician checked patient's understanding of advantages | 155 | 5.0 (4.0-5.0), 0-5 | 162 | 4.0 (3.0-5.0), 1-5 | 150 | .67 (.59 - .74*) |
| 5. Physician checked patient's understanding of disadvantages | 154 | 5.0 (4.0-5.0), 0-5 | 162 | 4.0 (3.0-5.0), 1-5 | 149 | .65 (.57 - .72*) |
| 6. Physician told how treatment options differ | 155 | 5.0 (3.0-5.0), 0-5 | 158 | 4.0 (3.0-5.0), 0-5 | 146 | .63 (.55 - .70*) |
| 7. Patient asked for clarification | 155 | 5.0 (3.0-5.0), 0-5 | 161 | 4.0 (2.0-5.0), 0-5 | 149 | .60 (.52 - .68) |
| 8. Physician said there is a choice | 156 | 5.0 (3.0-5.0), 0-5 | 160 | 4.0 (3.0-5.0), 0-5 | 149 | .65 (.57 - .72*) |
| 9. Physician said patient's opinion is important | 156 | 5.0 (4.0-5.0), 0-5 | 161 | 4.0 (3.0-5.0), 0-5 | 150 | .67 (.59 - .74*) |
| 10. Physician checked if he/she understood what is important for patient | 156 | 5.0 (4.0-5.0), 0-5 | 160 | 4.0 (3.0-5.0), 0-5 | 149 | .61 (.53 - .69) |
| 11. Physician helped patient weighing (dis)advantages | 156 | 5.0 (3.0-5.0), 0-5 | 161 | 4.0 (3.0-5.0), 0-5 | 150 | .63 (.55 - .70*) |
| 12. Physician gave patient time for weighing (dis)advantages | 156 | 5.0 (4.0-5.0), 0-5 | 161 | 4.0 (2.0-5.0), 0-5 | 150 | .55 (.47 - .64) |
| 13. Patient told physician what is important to him/her | 156 | 5.0 (4.0-5.0), 0-5 | 161 | 4.0 (3.0-5.0), 0-5 | 150 | .59 (.50 - .67) |
| 14. Patient weighed (dis)advantages~ | 155 | 5.0 (4.0-5.0), 0-5 | 160 | 4.0 (3.0-5.0), 0-5 | 148 | .59 (.51 - .68) |
| 15. Decision takes into account what is important for patient^ | 140 | 5.0 (5.0-5.0), 0-5 | 141 | 4.0 (4.0-5.0), 0-5 | 124<sup>-</sup> | .79 (.71* - .85*) |
| 16. Physician discussed what patient needs for weighing options^ | 16 | 4.0 (3.0-5.0), 0-5 | 19 | 4.0 (2.0-4.0), 1-5 | 7<sup>-</sup> | .43 (.12* - .71*) |

~Agreement was defined as the same item score or one point difference obtained by the iSHAREpatient and iSHAREphysician, since we considered it acceptable if scores from the respective viewpoints differed somewhat.
^We report item 15 if a patient/physician had reported that a decision had been made or if the response to that item was missing; we report item 16 if a patient/physician had reported that no decision had been made.
~We only report agreement if both patient and physician agreed on whether a decision had been made; a treatment decision was made according to both patient and physician for 124 decision making processes, while no treatment decision was made according to both patient and physician for seven decision making processes. For 10 decision making processes the patient indicated that a decision had been made, while no decision had been made according to the physician; it was the other way around for eight decision making processes. This was true also for 10 decision making processes according to the physician only, and for eight decision making processes according to the patient only.
~In our development article [14] we recommend researchers to split this item if they would like to determine whether weighing the options happened during or outside the consultation. In this study we presented these items at the end of the patient questionnaire: *I weighed up the advantages and disadvantages of the treatment options during the conversation*. Median (IQR): 4.0 (3.0-5.0) *I weighed up the advantages and disadvantages of the treatment options before or after the conversation*. Median (IQR): 4.0 (2.0-5.0).
\*Fleiss correction applied
CI = Confidence interval; IQR = interquartile range; P = proportion agreement

**Table 3.** Median and interquartile range for dimension and total scale scores of the iSHAREpatient (n=156 treatment decision making processes) and iSHAREphysician (n=162 treatment decision making processes), and for total and subscale scores of the comparison questionnaires

|  | Item | Patient |  | Physician |  |
| --- | --- | --- | --- | --- | --- |
|  |  | N* | Median (IQR) | N* | Median (IQR) |
| iSHARE dimension scores |  |  | **iSHAREpatient** |  | **iSHAREphysician** |
| 1. Choice awareness (0-5) | 8,9 | 156 | 5.0 (3.5-5.0) | 160 | 4.0 (3.0-5.0) |
| 2. Medical information (0-5) | 1-7 | 150 | 4.4 (3.6-5.0) | 158 | 3.9 (3.3-4.7) |
| 3. Preferences (0-5) | 10,13 | 156 | 5.0 (4.0-5.0) | 160 | 3.5 (2.5-4.5) |
| 4. Deliberation (0-5) | 11,14 | 155 | 5.0 (3.5-5.0) | 160 | 3.5 (3.0-4.5) |
| 5. Time for deliberation (0-5) | 12 | 156 | 5.0 (4.0-5.0) | 161 | 4.0 (2.0-5.0) |
| 6. Decision (0-5) | 15 or 16 | 156 | 5.0 (5.0-5.0) | 160 | 4.0 (3.0-5.0) |
| iSHARE total score (0-100) |  | 149 | 95.0 (77.1-99.5) | 155 | 75.0 (61.1-90.7) |
|  |  |  | **SDM-Q-9** |  | **SDM-Q-Doc** |
| SDM-Q (0-100) |  | 151 | 88.9 (71.1-97.8) | 161 | 77.8 (69.4-88.9) |
| COMRADE |  |  |  |  |  |
| Satisfaction with communication (0-100) |  | 130 | 72.0 (63.1-78.2) |  |  |
| Confidence in decision<sup>#</sup> (0-100) |  | 130 | 78.7 (71.0-79.3) |  |  |
| DCS<sup>#</sup> (0-100) |  | 149 | 15.6 (5.5-25.8) |  |  |
| Feeling uncertain<sup>#</sup> (0-100) |  | 153 | 16.7 (0.0-41.7) |  |  |
| Feeling uninformed (0-100) |  | 152 | 16.7 (0.0-25.0) |  |  |
| Feeling unclear about values (0-100) |  | 151 | 25.0 (0.0-33.3) |  |  |
| Feeling unsupported (0-100) |  | 152 | 8.3 (0.0-25.0) |  |  |
| Ineffective decision making (0-100) |  | 153 | 0.0 (0.0-12.5) |  |  |
| PEPPI-5 (5-25) |  | 155 | 24.0 (20.0-25.0) |  |  |

\*Numbers do not always add up to the total sample size, due to missing values.
<sup>#</sup>No a priori hypothesis was formulated regarding the correlation between this total or subscale score and either of the iSHARE questionnaires (Table 5); scores are reported for sake of information.
COMRADE = Combined Outcome Measure for Risk communication And treatment Decision making Effectiveness; DCS = Decisional Conflict Scale; IQR = interquartile range; PEPPI-5 = five-item
Perceived Efficacy in Patient-Physician Interactions; SDM-Q-9 = 9-item SDM-Questionnaire; SDM-Q-Doc = SDM-Questionnaire – physician version

**Figure 2.**
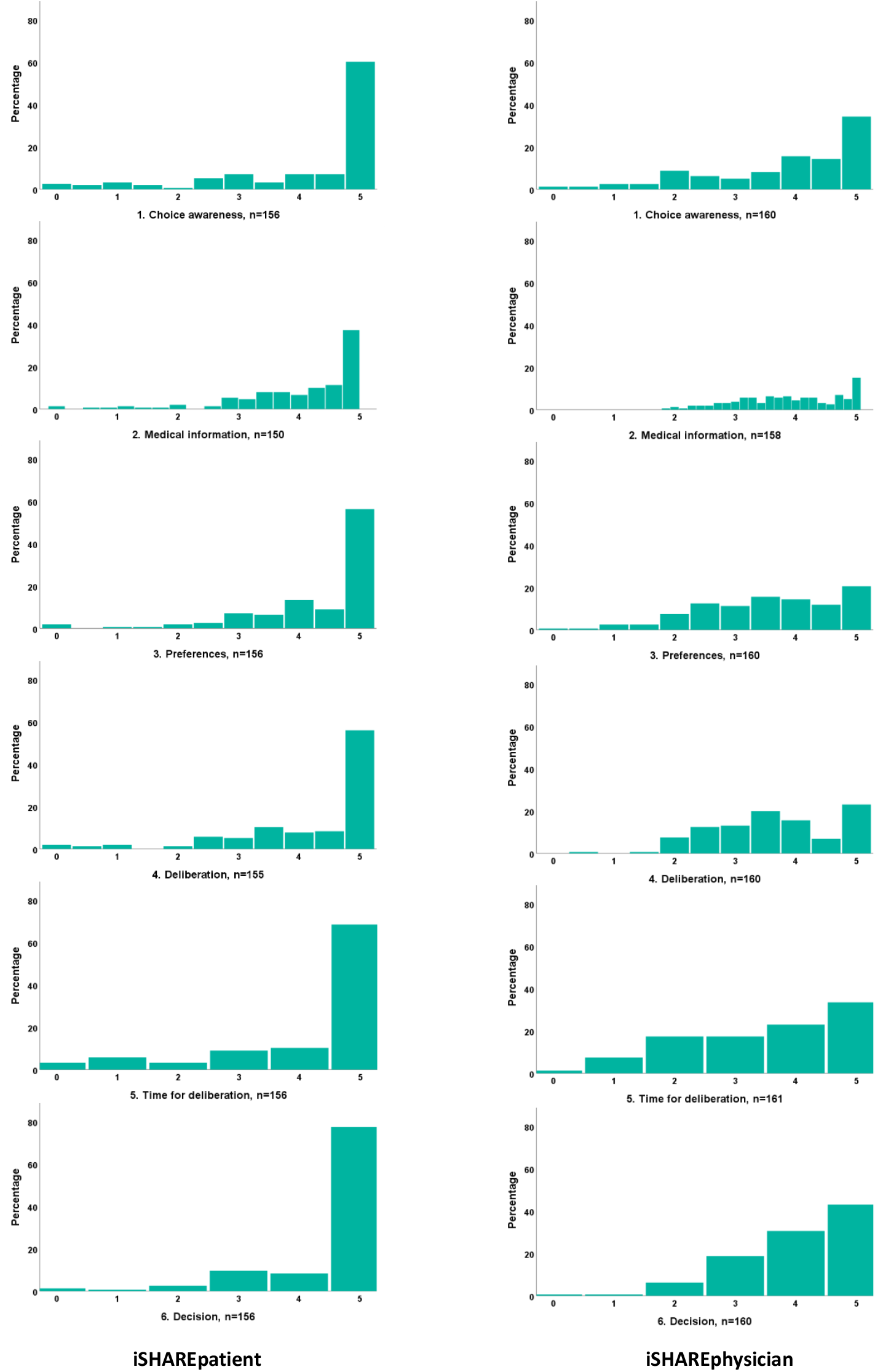
Dimension scores on the iSHARE questionnaires

### 3.3 Construct validity of the iSHARE questionnaires

Table 3 displays the median total and subscale scores on the comparison questionnaires used for hypotheses testing. The hypothesis formulated for the iSHAREphysician was confirmed. Nine out of ten hypotheses formulated for the iSHAREpatient were also confirmed (Table 5).

### 3.4 Test-retest agreement iSHAREpatient

In total, 112 patients completed the iSHAREpatient for the second time within 30 days post-consultation, of which 45 were excluded for various reasons (Figure 1). Mean time between test and retest was 11.1±3.7 (range, 4-24) days. Agreement at item level ranged from .55 (item 11) to .84 (item 15) (Table 4). Three patients had reported that no decision had been made at both test and retest and completed item 16 twice; agreement was .00. A post-hoc analysis in which we allowed item scores to differ one point, showed agreement ranging from .79 (item 7) to .97 (item 15) (Table 4).

**Table 4.** Test-retest agreement on item level for the iSHAREpatient (n=67 treatment decision making processes)

|  | Agreement<sup>~</sup> |  | Agreement<sup>-</sup> |
| --- | --- | --- | --- |
|  | N | P (95%CI) | P (95%CI) |
| 1. Physician explained advantages of treatment options | 67 | .64 (.52 - .74*) | .85 (.74* - .91*) |
| 2. Physician explained disadvantages of treatment options | 67 | .60 (.47 - .70*) | .88 (.77* - .93*) |
| 3. Physician explained (dis)advantages equally well | 67 | .64 (.52 - .74*) | .84 (.72* - .90*) |
| 4. Physician checked patient's understanding of advantages | 67 | .70 (.58 - .79*) | .91 (.81* - .95*) |
| 5. Physician checked patient's understanding of disadvantages | 66 | .61 (.48 - .71*) | .85 (.73* - .91*) |
| 6. Physician told how treatment options differ | 67 | .64 (.52 - .74*) | .84 (.72* - .90*) |
| 7. Patient asked for clarification | 67 | .60 (.47 - .70*) | .79 (.69 - .86*) |
| 8. Physician said there is a choice | 67 | .72 (.60 - .80*) | .87 (.76* - .92*) |
| 9. Physician said patient's opinion is important | 67 | .79 (.69 - .86*) | .93 (.83* - .96*) |
| 10. Physician checked if he/she understood what is important for patient | 67 | .69 (.57 - .78*) | .91 (.81* - .95*) |
| 11. Physician helped patient weighing (dis)advantages | 67 | .55 (.43 - .68) | .85 (.74* - .91*) |
| 12. Physician gave patient time for weighing (dis)advantages | 67 | .64 (.52 - .74*) | .91 (.81* - .95*) |
| 13. Patient told physician what is important to him/her | 67 | .76 (.65 - .84*) | .94 (.85* - .97*) |
| 14. Patient weighed (dis)advantages | 67 | .70 (.58 - .79*) | .91 (.81* - .95*) |
| 15. Decision takes into account what is important for patient<sup>^</sup> | 61 | .84 (.71* - .90*) | .97 (.88* - .98*) |
| 16. Physician discussed what patient needs for weighing options<sup>^</sup> | 3 | .00 (.03* - .56*) | .33 (.02* - .75*) |

<sup>~</sup>Agreement was defined as the same item score obtained both at test and retest.
<sup>-</sup>Agreement was defined as the same item score obtained both at test and retest, or one point difference as post-hoc analysis.
<sup>^</sup>We report item 15 if a patient had reported that a decision had been made or if the response to that item was missing; we report item 16 if a patient had reported that no decision had been made.
\*Fleiss correction applied
CI = Confidence interval; P = proportion agreement

### 3.5 Inter-rater agreement between the iSHAREpatient and iSHAREphysician

Inter-rater agreement between the iSHARE questionnaires ranged from .55 (item 12) to .79 (item 1 and 15). Seven patients and physicians both had reported that no decision had been made and completed item 16; agreement was .43 (Table 2).

## 4. Discussion and Conclusion

### 4.1. Discussion

In this study, we determined the measurement properties of the iSHAREpatient and the iSHAREphysician designed to assess SDM in oncology. As opposed to many existing questionnaires, the iSHARE questionnaires are based on a clear definition of the construct, provide a comprehensive assessment of the SDM process in- and outside consultations, and allow the assessment of both patient and physician behaviors [2, 14]. We have conducted a large-scale study, including patients and physicians from academic and non-academic hospitals, physicians from different specialties, patients with a variety of cancer diagnoses, and with treatment intents being either curative or palliative. The current analyses have shown high dimension and total scores on both iSHARE questionnaires, and good construct validity of the iSHARE questionnaires. The iSHAREpatient showed substantial test-retest agreement. Further, the iSHARE questionnaires correlated moderately with each other.

The iSHARE questionnaires, and especially the iSHAREpatient, showed high scores. More than 15% of the patients reported the highest possible score, which may be considered as a moderate ceiling effect [36]. Patient SDM questionnaires are known for ceiling effects. These may be caused by the so-called halo effect, leading people to unconsciously alter their judgement of others’ attributes based on their judgement of unrelated attributes [37]. To illustrate, if physicians are perceived to be friendly, the halo effect leads patients to evaluate their information-giving behaviors favorably instead of critically assessing them. Methods to reduce these effects, such as reflecting (stop-and-think) before rating the SDM process, have not been shown successful in patients [3]. We aimed to avoid ceiling effects by using an unbalanced response scale, that is, using a scale with more positively-labelled than negatively-labelled response options, thereby enabling more differentiation [21]. We further explicitly stated in the introduction of the iSHAREpatient that the questionnaire is not about satisfaction with the physician (Box 1) [14]. However, these precautions do not seem to have adequately addressed the problem. Of note, the high scores may have resulted from recruiting physicians from our network (i.e., researcher selection bias), some of whom had been trained in SDM and whose patients may actually have experienced high levels of SDM. Moreover, physicians may have, consciously or unconsciously, selectively approached patients with whom the decision making process was, or was expected to be, shared (i.e., physician selection bias). In addition, patients who declined participation may have been less involved in decision making (i.e., patient selection bias). A clear indication that our sample suffered from some form of selection bias were the remarkably high scores on the other questionnaires too. Two recent studies in Dutch cancer patients [38, 39] showed substantially lower SDM-Q-9 scores and higher decisional conflict scores. In addition, two recent studies in Dutch cancer patients [40] and Dutch cancer survivors [41] showed somewhat lower patients’ perceived self-efficacy compared to our sample. It is therefore important to await the scores in other samples before drawing definitive conclusions about the high scores. Of note, treatment decision making is often distributed across consultations and time [42] and half of the patients indeed thought about more than one consultation while completing the questionnaire.

The iSHARE questionnaires showed only very small numbers of missing values and no specific patterns, implicating acceptability of the items for both patients and physicians, and no systematic bias. Regardless, more research is needed on how to deal with missing values for instruments assessing formative constructs.

Our results demonstrated good construct validity (i.e., >75% of the results confirm our hypotheses [43]) of the iSHARE questionnaires. The iSHAREpatient and iSHAREphysician correlated highly (>.50) with the SDM-Q-9 and SDM-Q-Doc, indicating that the questionnaires measure the same construct [44]. The iSHARE questionnaires offer a more valid assessment of the SDM process since they cover both patient and physician behaviors. Hypotheses with regard to correlations with the COMRADE and DCS subscales were confirmed, adding to the proof for construct validity. Internal consistency of the DCS subscales seemed sub-optimal, a problem identified previously [45]. Further, two of three hypotheses regarding the PEPPI-5 were confirmed. To our knowledge no appropriate questionnaires were available at the time of designing the study for construct validity testing of any of the iSHAREphysician dimensions, nor for the Choice Awareness, Deliberation and Time for Deliberation dimensions of the iSHAREpatient. We recommend hypotheses testing for the other iSHARE dimensions once appropriate measurement instruments become available.

We determined test-retest agreement for the iSHAREpatient. This is a strength of the study, as this has not frequently been established for patient SDM questionnaires [2]. While several guidelines are available for kappa and intraclass correlations [43, 46], we are not aware of any criteria to label the proportion agreement. Using the labels proposed for the kappa [47], we propose that a proportion agreement of ≤30 is ‘slight’; >.30 ‘fair’; >.50 ‘moderate’; >.70 ‘substantial’, and >.90 ‘almost perfect’. This results in substantial agreement for four, moderate for eleven, and slight for one of the iSHAREpatient items. Higher agreement may be found if the period between the two assessments is even shorter. The time period should be long enough, so that participants will not remember their previous answers; yet patients risk forgetting about their and their physician’s behaviors if the period is too long. In addition, test-retest agreement of a questionnaire evaluating a decision making process may be different from one evaluating, e.g., a state such as quality of life, or an attitude. Consequently, we did a post-hoc analysis in which we allowed the item scores to differ one point; agreement was almost perfect for seven items, substantial for eight items and fair for one item. All in all, the results demonstrate substantial test-retest agreement.

We applied the same criteria to the agreement between the iSHAREpatient and iSHAREphysician scores, allowing one point difference; agreement was substantial for three, moderate for 12 and fair for one item, demonstrating moderate inter-rater agreement overall. As noted, some physicians had been trained in SDM and may have reflected more critically on the decision process than their patients. Patients’ and physicians’ ratings of communication, including SDM in oncology [24, 25] are known to correlate poorly, but it should be noted that correlations are not the appropriate measure for agreement [48, 49]. Only few studies calculated the kappa and proportion agreement [48]. To the best of our knowledge, we are the first to have calculated proportion agreement for patient and physician SDM scores in oncology, which makes it hard to compare results. We aimed to achieve good inter-rater agreement by using the same underlying construct for both questionnaires, using the same items and most importantly, extensively involving both patients and physicians throughout the development process of the questionnaires [14]. We recommend future users of the iSHARE questionnaires to consider which perspective is most feasible to determine or to use both, bearing in mind that they represent different perspectives.

The iSHARE questionnaires contain two versions of the last item; for the majority of decision making processes a decision had been made, so item 15 (The decision takes into account what is important for the patient) was reported. As a consequence there were not enough data to determine agreement for item 16 (The physician discussed what the patient needs to weigh the options). The iSHARE questionnaires may be applicable to healthcare settings outside of oncology, but we advise content validity testing first. We also recommend to determine cross-cultural validity when using the iSHARE questionnaires in languages other than Dutch. Finally, the findings should be considered in light of several limitations. As discussed, different forms of selection bias might have been present. Further, we aimed to include a broad range of patients, including in terms of education. Forty percent were highly educated, which may limit the representativeness of the sample for the patient population.

### 4.2. Conclusion

The iSHAREpatient and iSHAREphysician demonstrate good construct validity, substantial test-retest agreement (iSHAREpatient), and moderate inter-rater agreement. The dimension and total scores were high, which may have largely been caused by selection bias.

#### 4.3 Practice Implications

Results obtained using the iSHARE questionnaires provide information about the entire SDM process, about both patient and physician behaviors, from the perspective of patient and/or physician, and may be administered before or after the final decision has been made. The results may inform both physician-and patient-directed efforts to improve SDM in clinical practice, and dimension scores can be used to determine the impact of interventions or training on specific aspects of the SDM process.

## Data Availability

The Medical Ethical Committee of the Leiden University Medical Center (LUMC) approved the study (NL50551.058.14, P14.207), which was conducted according to the Dutch Medical Research Involving Human Subjects Act.

## ACKNOWLEDGEMENTS

We thank all participating cancer patients and physicians for their time and effort. We in particular thank the following physicians: Rob F.M. Bevers, MD, PhD; Patricia J.A.M. Brouwers, MD, PhD; Henk E. Codrington, MD; Ida E.M. Coremans, MD, PhD; Henk H. Hartgrink, MD, PhD; Fabian A. Holman, MD, PhD; Danny Houtsma, MD; Nils Knotter, MD; Roy F.P.M. Kruitwagen, MD, PhD; Saskia A.C. Luelmo, MD; Nikki D.M. van Luxemburg, MD; Rieneke. G. Moeri-Schimmel, MD; Femke P. Peters, MD, PhD; Inge R. Steenbakkers, MD; Willem H. Steup, MD, PhD; Bastiaan D.P. Ta, MD; Noortje Thielen, MD, PhD; Yvette M. van der Linden, MD, PhD; Erik J. van Gennep, MD; Evert J. Van Limbergen, MD, PhD; Arjan J. Verschoor, MD; Luuk N.A. Willems, MD, PhD. We thank Nanny van Duijn-Bakker (Department of Biomedical Data Sciences, Leiden University Medical Center, Leiden, The Netherlands) for her assistance in the data collection and Henrica C.W. de Vet, PhD for her advice regarding the calculation of agreement.

## Box 1.

iSHAREpatient† [14]

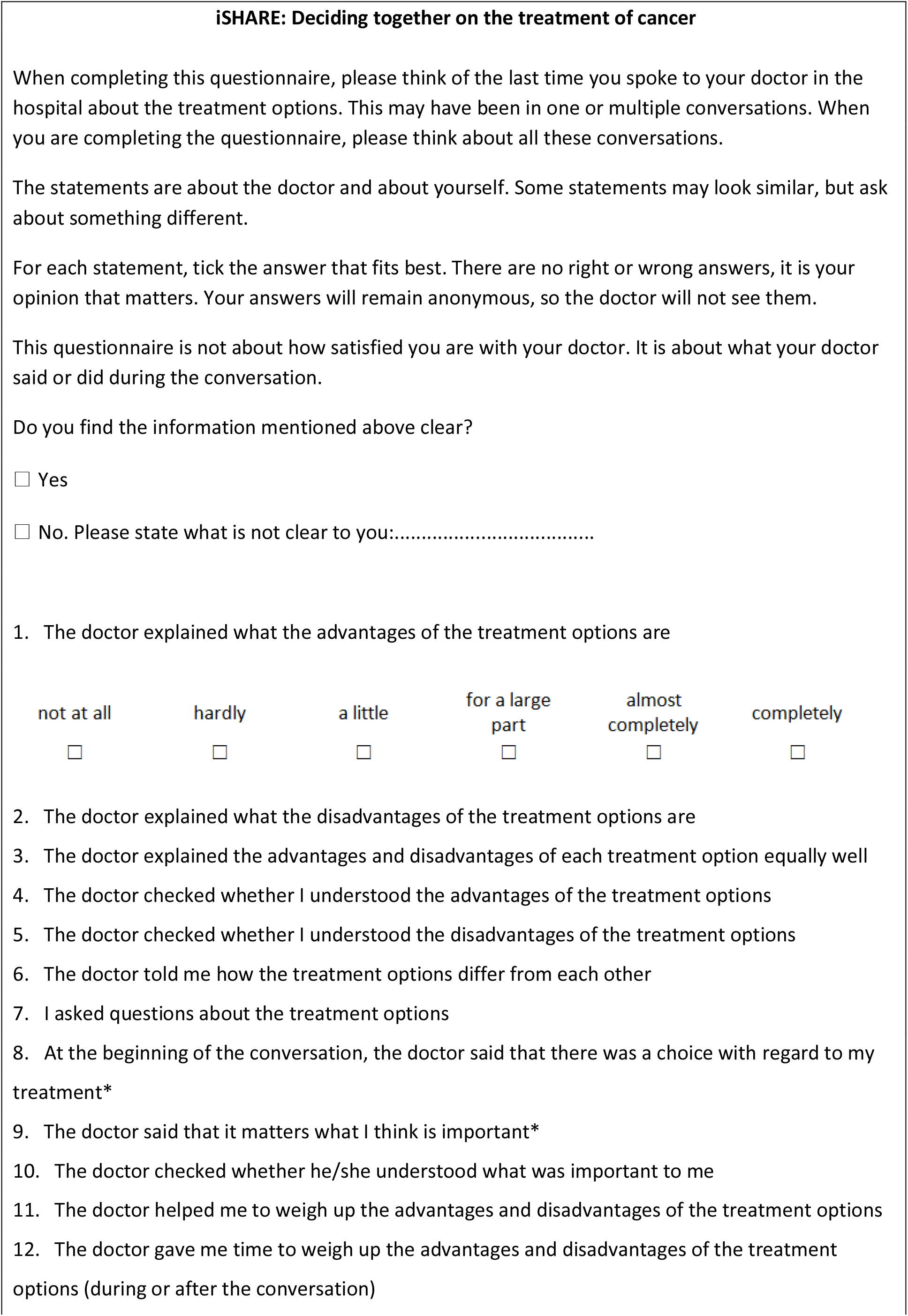
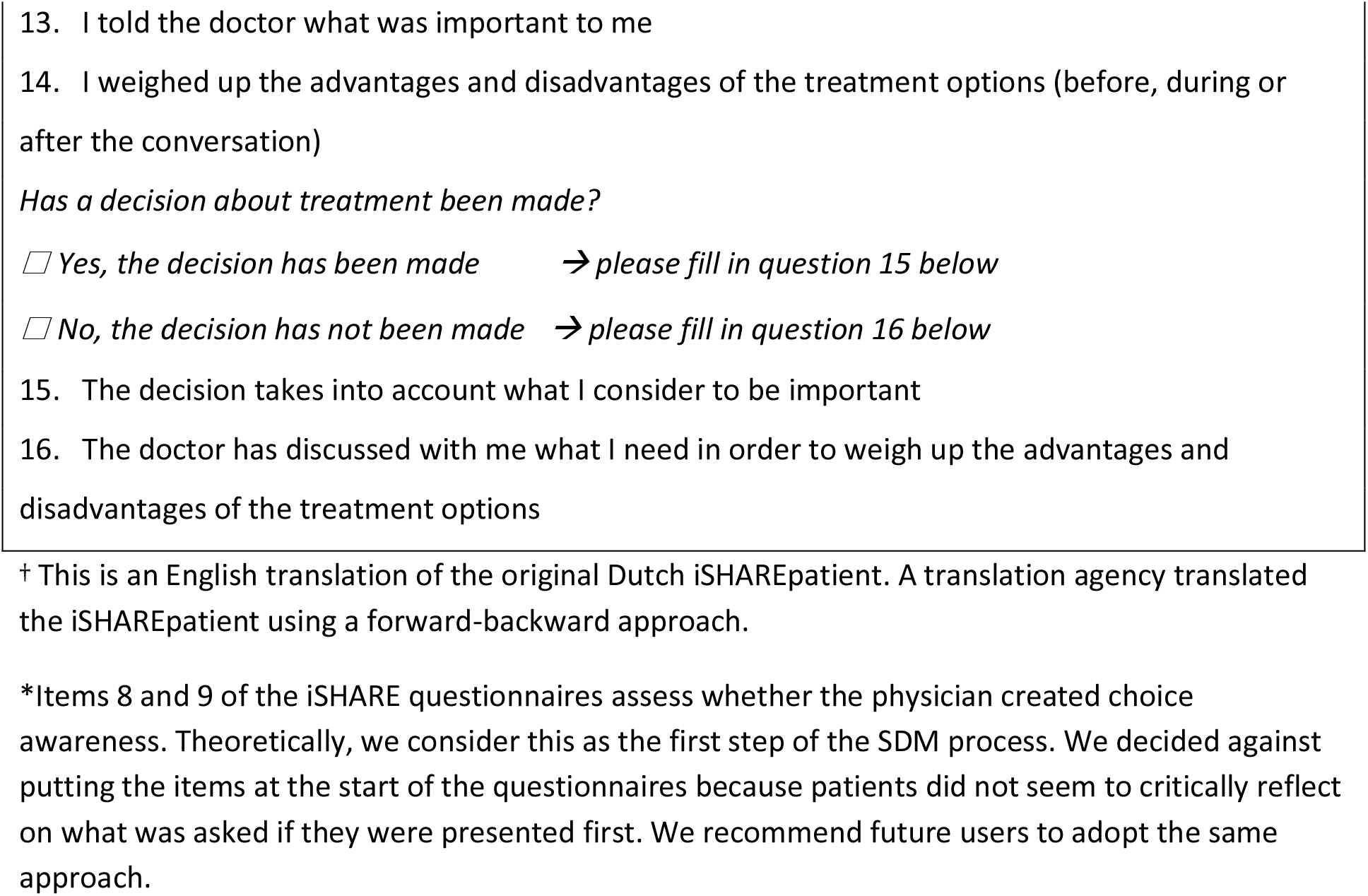

## Box 2.

iSHAREphysician† [14]

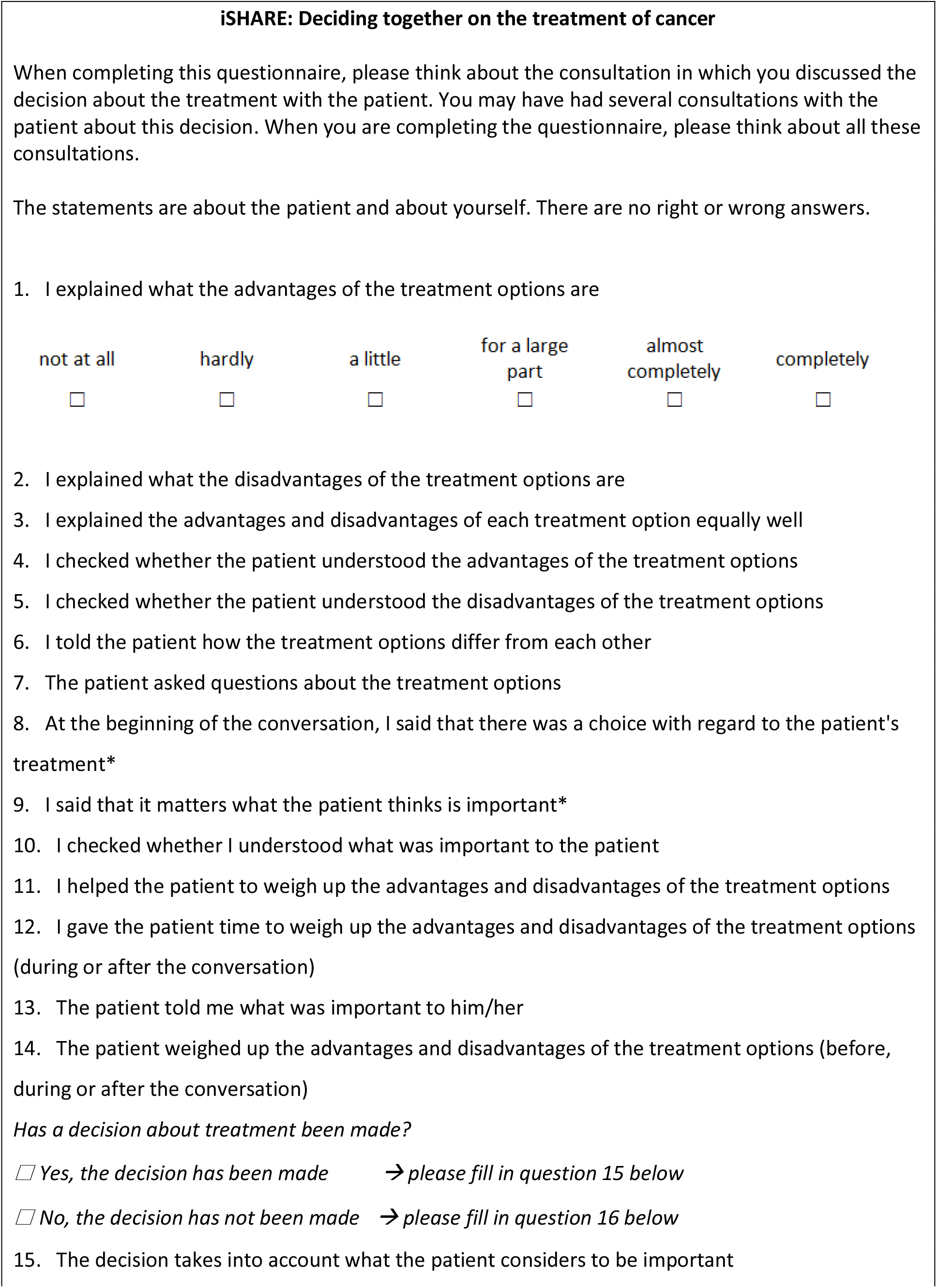
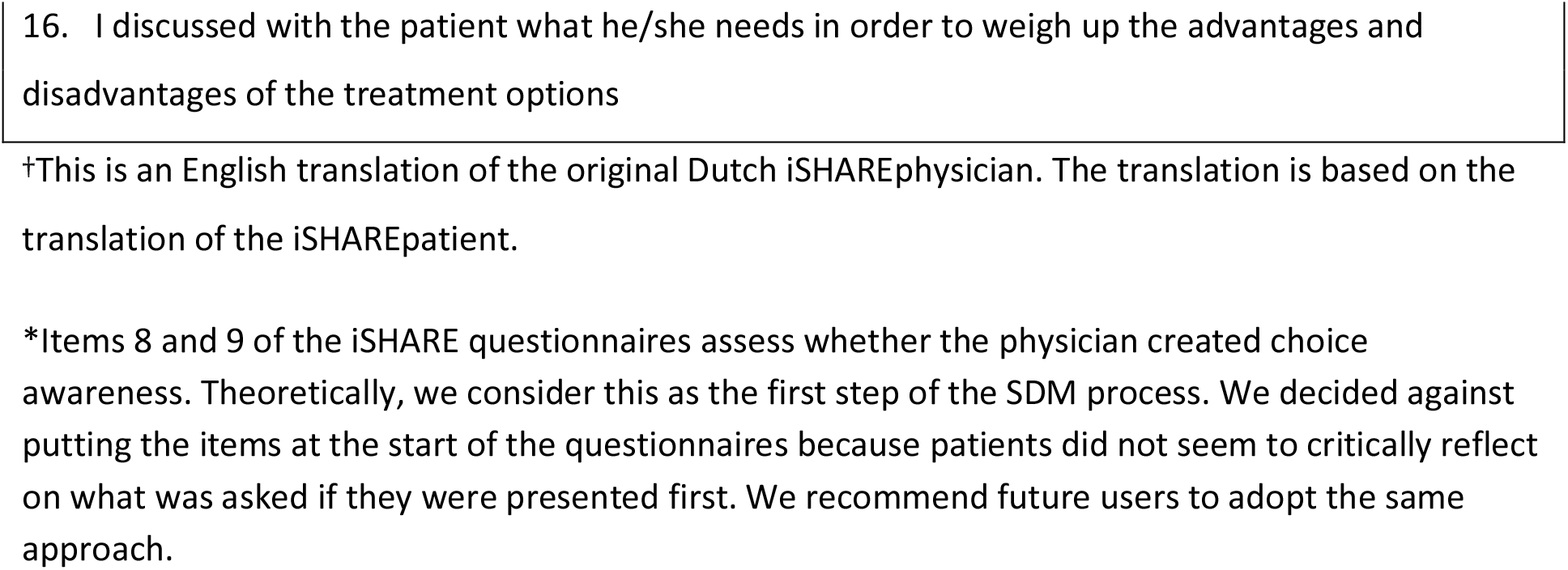

